# Increased Levels of N-Lactoylphenylalanine After Exercise are Related to Adipose Tissue Loss During Endurance Training in Humans With Overweight and Obesity

**DOI:** 10.1101/2022.09.07.22279536

**Authors:** Miriam Hoene, Xinjie Zhao, Jürgen Machann, Andreas L. Birkenfeld, Martin Heni, Andreas Peter, Andreas Niess, Anja Moller, Rainer Lehmann, Guowang Xu, Cora Weigert

## Abstract

**Objective:** The exercise-inducible metabolite N-Lactoylphenylalanine (Lac-Phe) has recently been shown to reduce food intake and adipose tissue mass in mice. We addressed whether Lac-Phe could have a similar function in humans.

**Methods:** Sedentary subjects with overweight and obesity completed an 8-week supervised endurance exercise intervention (n=22). Before and after the intervention, blood plasma levels of Lac-Phe were determined by UHPLC-MS in the resting state and immediately after an acute endurance exercise test. Adipose tissue and muscle volume were quantified by MRI.

**Results:** Acute exercise caused a pronounced increase in Lac-Phe, both before and after the intervention. Higher levels of Lac-Phe after acute exercise were associated with a greater reduction in abdominal subcutaneous and, to a lower degree, visceral adipose tissue during the intervention.

**Conclusions:** Lac-Phe produced during physical activity could boost weight loss in humans with obesity and overweight, possibly by transmitting or enhancing the appetite-suppressing effects of lactate. Exercise-induced Lac-Phe could be employed to predict and, potentially, improve the effectiveness of lifestyle interventions in subjects with overweight and obesity.

**Trial Registration:** Clinicaltrials.gov

NCT03151590

22 May 2017

https://clinicaltrials.gov/ct2/show/NCT03151590

## Introduction

The incidence of type 2 diabetes and related cardio-metabolic diseases is increasing worldwide. Physical activity and weight loss are two important pillars of diabetes prevention [1]. There is, however, a large variability in the effectiveness of exercise-based lifestyle interventions regarding therapeutic targets such as improvement of blood glucose control or reduction of adipose tissue mass that cannot satisfactorily be explained up to now [2–4].

The pronounced changes in skeletal muscle and whole-body energy metabolism induced by physical activity cause an increase in the concentration of a multitude of small metabolites in the circulation. A growing number of these metabolites have been found to not only reflect metabolic spillover but also play an essential role in mediating crosstalk between different cells and organs and thereby, the beneficial effects of physical activity [5–7].

N-Lactoylphenylalanine (Lac-Phe) is a pseudo-dipeptide generated from lactate and phenylalanine that has been reported to increase in a particularly pronounced fashion during and shortly after physical exercise [8,9]. A potential role in humans has not been reported yet, however, Lac-Phe has recently been shown to lower body weight and adipose tissue mass in mice rendered obese by a high-fat diet [9]. This prompted us to assess whether differences in the reduction in adipose tissue volume during an exercise intervention in humans with overweight and obesity could be attributed to differences in the exercise-induced production of Lac-Phe.

## Methods

### Study design and participants

All participants gave written informed consent. The study was approved by the ethics committee of the University of Tübingen and registered at Clinicaltrials.gov (NCT03151590). Details of the study protocol including recruitment and exclusion criteria have been published recently [10].

In brief, healthy subjects with <120 min of physical activity per week and a BMI >27 kg/m^2^ completed an 8-week supervised exercise intervention flanked by two acute exercise visits performed as follows: Blood was collected in the morning in the fasted state, 45 min before the commencement of exercise. In the meantime, the participants received a standardized breakfast (1 bun, 20 g butter, 1 slice of cheese, 150 g apple puree, water). Subsequently, they performed 30 min of bicycle ergometer exercise at the heartrate corresponding to 80% of their individual VO_2_peak. A second blood sample was collected 5 min after this bout of exercise. EDTA blood samples were immediately placed on ice, processed within 30 min and plasma stored at −80 °C.

The training intervention consisted of three times per week 1 hour supervised endurance exercise, 30 minutes each cycling and walking at 80% VO_2_peak. The target heartrate was maintained throughout the intervention. Determination of VO_2_peak and magnetic resonance imaging (MRI)-based quantification of adipose and lean tissue in arms and legs have been described [10,11]. Abdominal adipose tissue was segmented into visceral and non-visceral, which mainly consists of subcutaneous adipose tissue, using an automated procedure [11]. After excluding one subject with newly diagnosed autoimmune thyreoiditis, complete blood sample sets from 22 out of 26 subjects were available for metabolomics analysis.

### Quantification of plasma metabolites

The plasma levels of Lac-Phe, Lactate and Phe were determined using ultra high-performance liquid chromatography-mass spectrometry (UHPLC-MS). 50 μL of plasma were mixed with 250 μL of MeOH, vortexed 30 s and centrifuged for 20 min at 16,000 g, 4 °C. The supernatant was vacuum-dried in aliquots of 200 μl. Dry samples were resuspended in 50 μL 25% ACN/water. The analysis was performed on a Vanquish UHPLC coupled to a Q Exactive (both Thermo Fisher Scientific, Waltham, USA) operated in negative ion mode as previously described with slight modifications [12]. The separation was performed on a 2.1×100 mm ACQUITYTM UPLC HSS 1.8 μm T3 column (Waters, Milford, MA, USA). The mobile phases were (A) 6.5 mM ammonium bicarbonate in water and (B) 6.5 mM ammonium bicarbonate in 95% MeOH/water (B). The elution started with 2% B for 1 min, linearly changed to 100% B within 20 min, reverted back to 2% B, and equilibrated for 2.9 min (flow rate 0.35 mL/min, column temperature 50 °C). The Q Exactive was set to 140,000 resolution and full scan mode, mass scan range was 70-1050 *m/z*. Nitrogen sheath gas and nitrogen auxiliary gas were set at flow rates of 45 and 10 AU. Capillary and aux gas heater temperatures were 300 °C and 350 °C, respectively. The spray voltage was 3.00 kV. Parallel reaction monitoring was used to obtain high-resolution MS/MS spectra of Lac-Phe (*m/z* = 236.0928) with a resolution of 17500 and a collision energy of 30 eV. The internal standard d5-Phe (0.8 μg/mL in extraction solvent) was used to normalize signal intensities.

### Statistical analysis

Statistical analyses were performed using JMP 16 (SAS Institute Inc, Cary, North Carolina, USA). Longitudinal comparisons were performed using paired t-tests. Multiple linear regression analyses were performed on log-transformed data and adjusted for sex, age, baseline values of the respective tissue compartment or BMI, or change in muscle volume, as indicated. Normal distribution of the residuals was confirmed with the Shapiro-Wilk test in all analyses. A p-value < 0.05 was considered statistically significant.

## Results

Lac-Phe was identified by LC-MS/MS (Fig. 1A) and could be quantified in all samples (Fig. 1B). Acute exercise caused a significant increase in plasma Lac-Phe levels, both before and after the 8-week training intervention (Fig. 1B). Training had no effect on Lac-Phe concentrations in the resting state or after the acute bout of exercise (Fig. 1B), which was performed at the same relative intensity before and after the intervention. Plasma levels of Lac-Phe exhibited a significant correlation with phenylalanine (Fig. 1C, R^2^=0.35) and a very strong correlation with lactate (Fig. 1D, R^2^=0.82).

**Figure 1:**
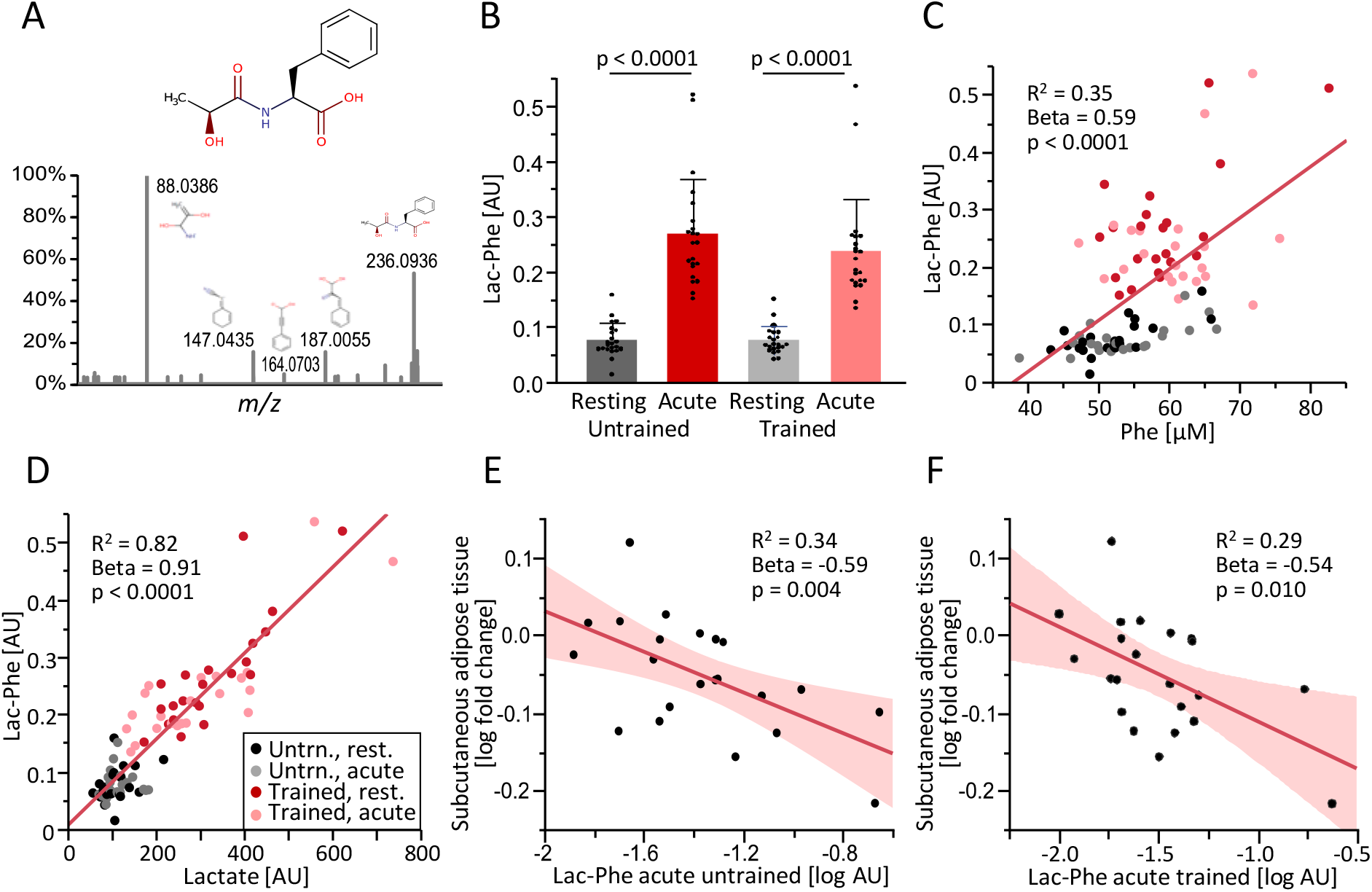
**A**: Structure of N-Lactoylphenylalanine (Lac-Phe) and identification by LC-MS/MS. **B**: Plasma levels of Lac-Phe in subjects with overweight and obesity (n=22) before (resting) or immediately after (acute) a 30-min endurance exercise session in the untrained state or after an 8-week exercise intervention (trained). **C-D**: Correlation of plasma Lac-Phe with plasma phenylalanine (**C**) and lactate (**D**). **E-F**: Correlation of the fold change in subcutaneous abdominal adipose tissue volume during the exercise intervention with the concentration of Lac-Phe after acute exercise before (**E**) and after the training intervention (**F**). R^2^, standardized beta coefficient (Beta) and p-value are shown for the simple linear regression (values from multiple regression presented in Table 2). AU, arbitrary units.

The training intervention resulted in an improvement in VO_2_peak and an increase in lean tissue, i.e. muscle mass, in the legs (Tab. 1). At the same time, BMI and abdominal subcutaneous and visceral adipose tissue were decreased (Tab. 1). The decrease in subcutaneous adipose tissue was inversely correlated to the plasma concentration of Lac-Phe after acute exercise, both before (Fig. 1E) and after the intervention (Fig. 1F) and also after adjustment for sex, age, and adipose tissue baseline values (results of multiple regression analyses shown in Tab. 2). The decrease in visceral adipose tissue was inversely correlated to the Lac-Phe concentration after the final acute exercise test and tended to be correlated after the first acute exercise test (Tab. 2).

**Table 1.** Anthropometric, fitness and metabolic data

|  | Untrained | Trained | p-value |
| --- | --- | --- | --- |
| Sex | 14 female / 8 male |  | - |
| Age [years] | 30 ± 8.9<br>(19 – 59) |  | - |
| VO<sub>2</sub>peak<sub>ergo</sub>/body mass<br>[mL/(kg*min)] | 25.0 ± 4.2<br>(18.3 – 32.3) | 26.5 ± 4.7<br>(16.0 – 34.9) | 0.042* |
| BMI [kg/m<sup>2</sup>] | 31.7 ± 4.5<br>(27.5 – 45.5) | 31.3 ± 4.7<br>(26.3 – 45.2) | 0.006* |
| Subcutaneous abdominal<br>adipose tissue [L] | 15.3 ± 5.9<br>(8.4 – 32.2) | 14.7 ± 6.1<br>(7.2 – 33.1) | 0.006* |
| Visceral adipose tissue [L] | 3.53 ± 1.65 (0.81<br>– 7.26) | 3.38 ± 1.57 (0.94<br>– 6.68) | 0.012* |
| Lean tissue legs [L] | 17.9 ± 4.1<br>(12.3 – 27.5) | 18.2 ± 3.9<br>(13.2 – 27.7) | 0.034* |
| Lean tissue arms [L] | 9.73 ± 1.96<br>(7.31 – 14.27) | 9.85 ± 2.26<br>(7.19 – 14.42) | 0.551 |
| Glucose fasting [mmol/L] | 5.09 ± 0.40 (4.61<br>– 6.00) | 5.02 ± 0.40 (4.33<br>– 5.61) | 0.336 |

N = 22, VO<sub>2</sub>peak N=21; mean ± SD (range of values); \* p < 0.05.

**Table 2.** Association of the increased Lac-Phe and lactate levels after acute exercise with the training response

| Training fold change | Lac-Phe<br>untrained acute | Lac-Phe<br>trained acute | Lactate<br>untrained acute | Lactate<br>trained acute |
| --- | --- | --- | --- | --- |
| Subcutaneous abdominal<br>adipose tissue [L] | Beta = -0.62<br>p = 0.004* | Beta = -0.52<br>p = 0.028* | Beta = -0.60<br>p = 0.008* | Beta = -0.39<br>p = 0.102 |
| Visceral adipose<br>tissue [L] | Beta = -0.42<br>p = 0.075 | Beta = -0.48<br>p = 0.037* | Beta = -0.23<br>p = 0.372 | Beta = -0.37<br>p = 0.123 |
| BMI [kg/m<sup>2</sup>] | Beta = -0.25<br>p = 0.279 | Beta = -0.15<br>p = 0.538 | Beta = -0.13<br>p = 0.600 | Beta = 0.07<br>p = 0.784 |
| Lean tissue<br>legs [L] | Beta = 0.37<br>p = 0.079 | Beta = 0.22<br>p = 0.357 | Beta = 0.42<br>p = 0.047* | Beta = 0.47<br>p = 0.036* |
| Lean tissue<br>arms [L] | Beta = 0.14<br>p = 0.584 | Beta = 0.08<br>p = 0.758 | Beta = 0.27<br>p = 0.279 | Beta = 0.08<br>p = 0.759 |

Multiple linear regression analyses with adjustments for sex, age and baseline values of the respective tissue compartment or BMI. N=22, Beta, standardized beta coefficient, \* p < 0.05.

Plasma lactate levels after acute exercise exhibited a similar, but slightly weaker correlation to the change in abdominal adipose tissue (Tab. 2). This association only reached statistical significance for the subcutaneous depot after the pre-training acute exercise bout but not for visceral adipose tissue (Tab. 2). Furthermore, lactate levels after acute exercise were positively correlated to the increase in muscle mass of the lower extremities during the intervention. Importantly, Lac-Phe levels after acute exercise still exhibited an inverse correlation to the change in subcutaneous adipose tissue when additionally adjusting for the change in leg muscle volume in the multiple linear regression models (p=0.020, standardized Beta coefficient=-0.56 for the pre-training and a trend of p=0.088, Beta=-0.42 for the post-training exercise bout).

No significant correlation of Lac-Phe or lactate could be observed with the changes in BMI or in the lean tissue of the arms (Tab. 2). As expected, the latter was not increased by the training scheme, i.e. by cycling and treadmill exercise (Tab. 1).

## Discussion

The metabolite Lac-Phe is produced during physical exercise and has recently gained attention as a mediator of weight loss in mice [9] but its relevance and function in humans remain to be demonstrated. We provide a first clue by showing that higher levels of Lac-Phe after exercise are related to a greater reduction in abdominal subcutaneous and, to a lower extent, visceral adipose tissue in subjects with obesity during a supervised exercise intervention.

Lac-Phe produced during physical exercise has been shown to reduce obesity by lowering food intake in mice [4]. Food intake has also been identified as an important factor for weight loss in humans with obesity [13,14]. While food intake has not been systematically assessed in our study, it seems plausible that higher levels of Lac-Phe did cause a greater transient suppression of hunger after each exercise session that contributed to a negative energy balance.

A reduction of food intake following physical exercise may seem counter-intuitive at first sight. However, in humans, a compensatory increase in calorie intake only occurs with a delay in the range of days and more pronouncedly in lean subjects [15] while in subjects with obesity, physical activity may even reduce food intake by restoring the physiologic regulatory circuitry [13,14]. Acutely, physical activity causes a repression of hunger that serves the purpose of preserving blood flow to skeletal muscle and that correlates with the circulating concentrations of lactate [16]. Studies of lactate administration have supported an appetite-suppressing effect of this metabolite and suggested different potential mechanisms of action [16,17]. Since lactate drives the formation of Lac-Phe [8], which then peaks after lactate [9], it could be speculated that Lac-Phe mediates a more sustained appetite-suppressing signal than lactate itself. One mode of action for Lac-Phe could be via G protein-coupled receptors in the brain, the central regulator of food intake [18,19].

Independent of its signalling function, Lac-Phe could serve as a biomarker to predict and, potentially, optimize the individual response to exercise-based lifestyle interventions. This is particularly relevant given the large variability in the extent to which different subjects benefit from a given exercise scheme [2–4]. Exercise intensity is usually personalized, e.g. based on the individual VO_2_peak as in our study, but parameters to determine the ideal intensity or modality of exercise that are most suitable for an individual have not been available up to now. Lac-Phe could serve as such a parameter since subjects exhibiting higher levels after an exercise test achieved a greater reduction in adipose tissue mass despite having exercised at the same relative VO_2_peak. In addition to modifying the exercise scheme, future studies could also aim at boosting the Lac-Phe response in order to enhance weight loss and thereby prevent type 2 diabetes and related cardiometabolic diseases.

## Data Availability

The data will only be made available to interested researchers upon reasonable request to such a degree as privacy and consent of the study participants is not compromised.

## Abbreviations

ACN: acetonitrile
BMI: body mass index
MeOH: methanol
MRI: magnetic resonance imaging
UHPLC-MS: ultra high-performance liquid chromatography-mass spectrometry.

## Acknowledgements

The manuscript has been published as a preprint (DOI: 10.1101/2022.09.07.22279536).

## Ethics Approval and Consent to Participate

All participants gave written informed consent. The study was approved by the ethics committee of the University of Tübingen and registered at Clinicaltrials.gov (NCT03151590).

## Author Contributions

MHo analyzed and interpreted the data and wrote and edited the manuscript. XZ performed LC-MS analyses. JM performed whole-body MRI. ALB, MHe, AP and, AN provided scientific guidance and contributed to the discussion. AM designed the study and analyzed anthropometric data. RL provided scientific guidance and experimental design and contributed to the discussion. GX provided scientific guidance and experimental design, contributed to the discussion and reviewed the manuscript. CW designed the study, supervised the whole project, contributed to the discussion, reviewed the manuscript and is the guarantor of the study. All authors approved the final version of the manuscript.

## Data Availability Statement

The data will only be made available to interested researchers upon reasonable request as far as privacy and consent of research participants are not compromised.

